# High Sensitivity of the BioNote Anigen™ Rapid Rabies Antigen Test in Detection of Diverse Rabies Virus Variants Suggests Utility for Global Rabies Control

**DOI:** 10.1101/2025.10.25.25338785

**Authors:** Lillian A. Orciari, Vaughn Wicker, Sarah Bonaparte, Pamela A. Yager, Claire Hartloge, Kirk Silas, Melanie Seiders, Puja Patel, Annette Regec, Miguel Maldonado-Cedeño, Ramon F. Flores Ramos, Sharon Messenger, Ruth Lopez, Christopher Preas, Alice Price, Kimberlee B. Beckmen, Kristin E. Campbell, Melanie Goff, Chis Vogt, Lisa Wingerter, Kristie L. Schwarzkopf, Yimer Mulugeta, Dessalegn S. Fujaga, Teresa Fields, George Dautu, Pierre Dilius, Crystal Gigante, Edgar Condori, Christina L. Hutson, Panayampalli S. Satheshkumar, Ryan M. Wallace

## Abstract

Point-of-care, immunochromatographic tests have not reliably detected rabies virus and have not utilized a consistent diagnostic protocol. International diagnostic standards established by the World Organization for Animal Health require strategic and large-scale validation studies prior to widescale use. The United States (US) Centers for Disease Control and Prevention, National Rabies Reference Laboratory undertook a large-scale validation of the BioNote Anigen™ Rapid Rabies Antigen Test in collaboration with eight US public health laboratories and three international laboratories. Modifications were made to manufacturer instructions to maximize antigen concentration in the test diluent. A total of 1,399 samples underwent paired testing with one of three US gold standard tests, consisting of 31 types of animals and 9 rabies virus variants. Test sensitivity and specificity was 97.11% (CI: 95.21% - 98.27%) and 99.89% (CI: 99.36% - 99.98%). Fourteen samples resulted in false-negative results, primarily impacting dogs that were euthanized early or shown to have a low viral load. Limit of detection studies found that false-negative results often occurred when the sample had a PCR Ct value ≥23. The BioNote Anigen™ Test performed well across diverse RVVs found in North America. Sensitivity of the test was slightly lower than, but not statistically inferior to, the minimum 98% value established for the current gold-standard tests. This is the largest systematic evaluation of a rabies point-of-care test that includes diverse RVV and animal type and results suggest that the BioNote Anigen™ Test with the procedural changes would have broad benefits for rapid diagnosis in animals.

## Introduction

Rabies virus has a unique viral pathogenesis that relies upon neurotropic centripetal movement from the site of inoculation to the central nervous system[1, 2]. Exponential proliferation within neurons results in neurologic decline of the animal (or human) and display of rabies clinical signs [3, 4]. Infected mammals are not infectious until the virus reaches the central nervous system and then migrates through cranial nerves to the salivary glands. From the time of infection through the time of central nervous system infection, there are no diagnostic methods that can reliably and consistently detect the virus, limiting timing and approaches to diagnosis [5-8].

In humans, ante-mortem (AM) rabies diagnostic approaches are available; however, these require invasive sampling methods (saliva, serum, cerebral spinal fluid, and nuchal skin biopsy) and advanced diagnostic testing capacities (rabies virus neutralizing antibody detection, indirect fluorescent antibody detection, antigen detection, and nucleic acid detection) [6, 9, 10]. Very few laboratories maintain this capacity, even in the United States [11]. AM testing is not well-studied in animals and is not recommended by international agencies such as the World Organization for Animal Health (WOAH) and the World Health Organization (WHO) [6, 8]. Animals, alive or deceased, suspected of having a rabies virus infection should undergo post-mortem (PM) testing. Unlike the less invasive AM procedures, PM testing requires a necropsy/autopsy to obtain full cross-section of the brain stem and representative aliquots of the cerebellum, which are then tested by WOAH/WHO recognized methods [Direct Fluorescent Antibody Test (DFA), Direct Rapid Immunohistochemistry Test (DRIT)] or nucleic acid detection methods [Real-time Reverse Transcription -Polymerase Chain Reaction Test (RT RT-PCR)]. When utilizing gold-standard methods, diagnostic sensitivity and specificity should exceed 98% [6]

Despite more than half a century of highly accurate rabies diagnostic methods, a global landscape analysis conducted in 2021 found that nearly all countries in Africa and Asia have inadequate surveillance and testing programs [12]. This dearth of surveillance and testing has been identified as a leading factor for rabies’ longstanding status as a neglected disease. Studies have consistently identified barriers with current diagnostic approaches which render them unlikely to be utilized effectively in low- and middle-income countries, where an estimated 70,000 people die from the disease each year. Diagnostic barriers include a lack of the following: trained personnel to collect samples, sample transportation networks to centralized labs, rigorous technical training (relegating the testing to only centralized labs), cost of and access to reagents (often >$30 per sample tested), and feedback mechanisms to ensure test results are shared back to the community in which the animal originated [13].

Point-of-care tests (also referred to as rapid tests, lateral flow tests, lateral flow devices, immunochromatographic assays, or virus antigen tests) have long been a priority and a challenge for the rabies community [14]. Expert WOAH evaluations have consistently found prospective commercial tests to be unreliable [15-26]. Additionally, many of these test kit manufacturers advise on the collection and testing of specimen-types that have extremely low sensitivity (e.g. saliva), even among gold standard diagnostic tests. Since 2020, one commercially available point-of-care test has undergone multiple small-scale, field-based evaluations that have suggested the device may have high sensitivity and specificity for rabies virus antigen in brain tissues, primarily in dog rabies endemic areas. While encouraging, these studies are not adequate in design or scale to satisfy international standards to validate a new diagnostic method [Figure 1, 15, 18, 19, 23, 25-27].

**Figure 1:**
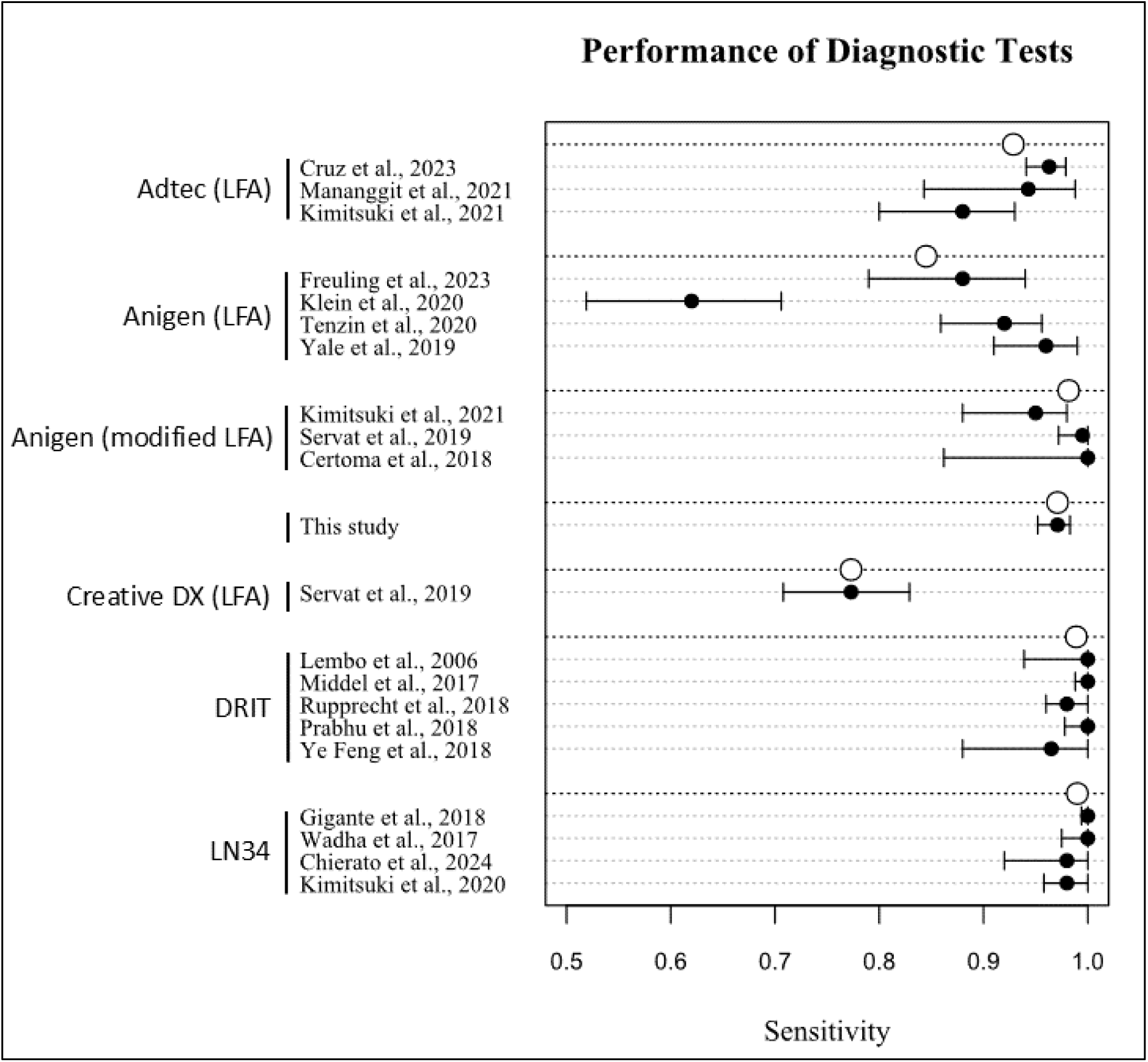
Previous Evaluations of Gold Standard Rabies Assays to the BioNote Anigen Test.

The U.S. Centers for Disease Control and Prevention (CDC) is a WOAH Reference Laboratory for Rabies, WHO Collaborating Center for Rabies, and serves as the U.S. National Rabies Reference Laboratory (NRRL) [28, 29]. Under CDC’s Terms of Reference, CDC has a mandate to support the evaluation and validation of rabies diagnostic tests. CDC developed the first rabies-specific antigen detection test, the DFA test, in 1958 and later established strict guidelines for performance of the National Standard Protocol for Rabies Diagnosis by DFA in the U.S. (2003) [30]. The DFA has been an international gold standard test for more than six decades and is still a primary diagnostic test for most of the world [31]. CDC developed the direct rapid immunohistochemical test (DRIT) in 2004 for antigen detection using only a standard light microscope [32, 33]. The DRIT was recognized as a standard for rabies diagnosis equivalent to DFA by WHO and WOAH in 2013 [6, 8]. In 2018, CDC developed and validated a novel molecular method for rabies diagnosis by real-time RT-PCR (LN34 Assay) [6, 34-37]. In 2020, the LN34 Assay was the first molecular method to be recognized as a gold standard test for rabies diagnosis in animals and humans by WHO and WOAH. Following these processes that led to successful recognition of numerous gold standard diagnostic methods, CDC undertook a large-scale evaluation of the BioNote Rapid Rabies Anigen Test, an immunochromatographic lateral flow assay (LFA), to inform international agencies on its recommended uses.

## Methods

CDC’s NRRL conducted a methodologic review of the BioNote package insert prior to initiating this evaluation with the purpose of identifying pre-validation study modifications to maximize sensitivity of the kit [27]. Several necessary modifications were identified based on knowledge of rabies pathogenesis, diagnostic experience in the distribution of rabies antigens, and experience in immunochromatographic testing methods. Notably, these modifications included: (a) requiring the exclusive testing of a full cross section of brainstem and not diluting this tissue with other tissues (cerebellum and hippocampus) which are less likely to contain rabies antigen, (b) that the brain stem tissue should not be diluted to 10% suspension, as is the manufacturer instructions, (c) that the brain stem be minced into a fine paste to increase access to antigen, (d) the transferal of minced brain paste using the swab provided by the kit to an empty 2 mL tube, (e) gently rubbing the swab against the tube sides to coat the swab with brain and remove excess tissue conglomerates, (f) transferal of the brain coated swab to the tube of diluent provided in the kit, and <u>slowly</u> blend brain into solution to avoid production of an emulsion with clumps. This study did not evaluate the alternative brain tissues and saliva samples recommended in the package insert which are not in alignment with international standards for rabies testing and should not be utilized [6, 8]. A CDC modified protocol was developed and shared with all participating laboratories as well as either an on-site or virtual training to ensure methodologic consistency (Appendix A). Five BioNote Rapid Rabies Ag Test kit lots: 18010006, Code: DEN (Exp. Date Feb. 28, 2022), 1801D011, Code: DEN (Exp. Date Jun. 20, 2023), 1801 D015, Code: DENN (Exp. Date May, 08, 2024, 1801D019, Code: DENN (Exp. Date Jun 25, 2025), 1801D031, Code: DENN (Exp. Date Aug. 25, 2026) were evaluated in this study.

Participating laboratories were selected based on their roles as state public health or veterinary rabies laboratories which receive and test through routine test submissions the diversity of rabies virus variants (RVV) and host species present in the United States [29, 38]. Six public health laboratories in states Arizona, California, Illinois (Springfield and Carbondale), North Dakota, Pennsylvania and the territory of Puerto Rico and Alaska Department of Fish and Game (AK DFG) were recruited as test sites, in addition to the CDC NRRL. Besides the above-mentioned laboratories participating as kit testing sites, the Texas and Kentucky State Public Laboratories provided both DFA positive and negative samples to fill gaps in species or variant sample numbers, which were tested at the CDC NRRL. Three international field sites (Ethiopia, Zambia and Haiti) provided test validation data for comparison with DFA (Ethiopia and Zambia) and real-time RT-PCR at CDC (Haiti), to represent dog-maintained RVV which have been eliminated from the United States.

All sample testing occurred through routine public health and animal health rabies diagnostic pathways, on animals suspected of being infected with rabies virus; no animals were euthanized or tested for the sole purpose of this validation study [29]. All participating laboratories utilized a gold-standard (GS) primary test (recognized by WHO, WOAH or US Council for State and Territorial Epidemiologists), either Direct Fluorescent Antibody (DFA) test, LN34 Real-Time reverse transcription Polymerase Chain Reaction (RT-PCR), or the Direct Rapid Immunohistochemical Test (DRIT) [39]. However, primary testing was performed by the six public health labs by strict adherence to the National Standard Protocol for DFA,. Initially, the AK DFG used the DRIT, as the GS for primary testing since it was that lab’s standard operating procedure (SOP) for surveillance samples. However, aliquots were stored in TRIzol™ reagent [to inactivate potential endemic highly pathogenic avian influenza virus (HPAIV) detected in suspect AK rabies samples] prior to shipment to CDC for confirmatory rabies testing, or due to the unavailability of DRIT reagents, GS testing by real-time RT-PCR only (no tissues available for DFA). Aliquots of tissue were separately processed through the BioNote Anigen test. Sample information was collected on a standard form, which included the state and county of sample origin, testing laboratory, type of animal, sample condition, gold-standard test result, a description of the gold test result (e.g. DFA Intensity and Distribution or PCR Cycle Threshold (Ct) value), LFA test result, LFA band strength (qualitative), and the rabies virus variant (if applicable). Discordant samples were submitted to the CDC NRRL for further testing and included in the validation study if they met routine criteria for reporting definitive test results.

### LN-34 Assay and Sequence Analysis Sub-Study

In addition to routine use for determination of the RVV of the samples within the study, further analysis was performed for 3 out of the 14 brain tissues that produced false-negative results with the BN-LFA (BN-LFA) to determine if there were distinct differences in the viral nucleoprotein gene sequences which translated, and could explain the lack of antigen (rabies N-protein) detection. Total RABV RNA was extracted from 3 brain representative brain samples using the Direct-zol RNA miniprep kit (R2051 Zymo, Irvine, CA, USA) details of molecular testing can be found at https://www.protocols.io/private/5c970341ebdf05cba17e58ebc16dff08.

The real-time RT-PCR LN34 assay was used to confirm RABV RNA in the negative samples mentioned above [37]. The complete N gene was amplified using primers previously described [40] and SuperScript™ IV One-Step RT-PCR System (12594100 Invitrogen). A 20µl RT-PCR reaction mix that contained 4 µl DNase/RNase-free water, 0.5 µl of forward and reverse primers (20 µM), 5 µl RNA, and 10 µl 2X Platinum SuperFi RT-PCR Master Mix with 0.2 µl SuperScript IV RT Mix were run on a thermal cycler using the following conditions: 50°C for 10 min, 98°C for 2 min; 35 cycles of 98°C for 30 sec, 56°C for 30 sec, 68°C for 2 min 30 sec; final extension at 72°C for 5 min. N gene amplicons were purified using ExoSAP-IT ™ Express PCR Product Cleanup Reagent (Applied biosystems, Waltham, MA). 5 µl of purified amplicons was added to the reaction mix containing 2.3 µl of DNase/RNase-free water, 10 µl of 2x GC buffer I, 2 µl of dNTPs, 0.2 µl of Takara LA Taq, and 0.5 µl of unique barcoded primer from Expansion 1-96 EXP-PBC096 (ONT, Oxford, UK). The reaction mix was run on a thermal cycler at 94°C for 1 min; 14 cycles of 94°C for 30 sec, 62°C for 30 sec, 72°C for 2 min; final extension at 72°C for 5 min.

DNA concentration of each sample was quantified using the Qubit dsDNA HS (High Sensitivity) assay (Invitrogen, Waltham, MA), and 200 ng of each amplicon was pooled and purified with 0.65X AMPure XP beads (Beckman, Brea, CA). Sequencing library was prepared using 500 ng of the pooled DNA and the ligation sequencing SQK-LSK109 kit (ONT, Oxford, UK). The library was sequenced using a flow cell FLG001on a MinION MK1B. Previously described script were used for base calling and demultiplexing [40]. Final consensus sequences were aligned to SAD B19 RABV (GenBank M31046) and edited manually in Bioedit v7.2.5.

### Dilution Study

Brain samples from a raccoon infected with Eastern Raccoon RVV (assay sensitivity 99.4%), a Texas skunk infected with South-Central Skunk RVV (assay sensitivity 92%), and a horse infected with PR Mongoose RVV (assay sensitivity 100%) with variable antigen distribution were chosen to determine if there were differences in the affinity of the antibodies within the BioNote test cassette for these divergent rabies virus variants. From those samples, approximate 100 mg cross-sections of brain stem were homogenized in 900 mL PBS. Five additional serial 10-fold dilutions were made from the original 10% (0) suspension in PBS representing 1:10 to 1:100,000 dilutions. 100 µl of the original sample suspension and 100 µl of each of the subsequent 10-fold dilutions made in PBS were mixed by vortexing and added to 1 mL of Trizol and the premeasured BioNote sample diluent provided with the kit and RNA extraction followed by real-time RT-PCR respectively. LFA was performed on the dilution series and analyzed as described in this study.

RNA extraction was completed using the Direct-zol RNA miniprep kit (R2051 Zymo, Irvine, CA, USA) as described previously. Real-time RT-PCR was completed and analyzed as described previously using the LN34 to detect Lyssavirus RNA [34, 37]. In brain tissue samples, a positive result in the LN34 assay is defined by a Ct value below or equal to 35. The diagnostic threshold is shown by a horizontal line in figure 3. All values above 35 are inconclusive (Ct 35.01 – 39.99) or undetected by the assay (Ct 40). The graph in figure 3 was constructed in R (version 4.4.0) using the ggplot2 package.

### Data Analysis

Data were submitted to the CDC NRRL and stored in Microsoft Excel. Data cleaning comprised of standardizing nomenclature and spelling of animal type, animal common names, and RVV. Samples were censored from analysis under the following conditions: (a) if the sample was noted to not comply with testing requirements (typically lack of full cross section of the brainstem), (b) samples that failed to produce a control band on the LFA, (c) samples from rabbits, which have been demonstrated to consistently produce false-positive LFA reactions. In several instances, where two gold-standard tests were conducted and produced inconsistent results, if either were reported as positive, then it was considered positive for the purposes of analysis, as would be standard procedure in a public health rabies laboratory. All samples were assigned to a rabies virus variant territory through a tiered decision algorithm. Priority 1 samples were those where variant testing was conducted and reported and were categorized based on these test results. Priority 2 samples were bats that lacked variant testing and were categorized as bat variant. Priority 3 samples were foxes from Arizona that lacked variant results and were categorized as Arizona Gray Fox Variant. Skunks from Arizona without variant results were categorized as South-Central Skunk Variant. All other terrestrial animals from Arizona were placed in both categories for analyses related to variant, as both RVV are endemic in the state [29]. Priority 4 samples included all other terrestrial animals without variant results and were placed in the variant category that predominates in the county of origin [38].

A descriptive analytic approach was taken to characterize the range of sensitivity and specificity of the LFA across a broad range of mammalian animals and RVVs. LFA sensitivity was defined as the number of samples positive by both the gold standard and LFA (True Positives), divided by the sum of these true positive and samples that were positive by the gold standard but negative by the LFA (False-negatives). Specificity was defined as number of samples that were negative by both the gold standard test and LFA (True Negatives) by the sum of these true negatives and samples that were negative by the gold standard but positive by the LFA (False Positives). Confidence intervals for sensitivity and specificity of LFAs were calculated using the Wilson score method. As LFA band strength has been a notable concern for these devices, qualitatively reported LFA band strength, categorized as “strong”, “normal”, “weak”, or “absent”, were compared by animal type and RVV to the reaction strength of the paired gold standard test [30, 33, 37]. Additional information was collected and analyzed for false-negative LFA specimens, including: (a) Breed (b) Age at Death, (c) Day of Death Post-Symptom Onset, (d) Vaccination History. Where noted, the Fisher’s Exact Test for sensitivity was applied to determine if significant differences in sensitivity occurred between comparison groups.

## Results

Eight U.S.-based rabies diagnostic laboratories, CDC, and three international laboratories contributed 1,399 LFA results to this evaluation, of which 1,362 (97%) met inclusion criteria (Table 1, Figure 2). Thirty-seven specimens were ineligible: 15 due to failure to produce a definitive result from the gold standard test, 12 due to failing to produce a definitive result from the BN-LFA, and 10 due to inadequate sample quality. Among the 1,362 specimens meeting inclusion criteria, 877 (64%) were definitively negative for rabies virus and 485 (36%) were definitively positive. Of 485 true positive specimens, 14 (2.9%) were negative by the BN-LFA, resulting in a sensitivity of 97.1% (CI: 95.2% - 98.3%; Table 1). Among 877 true negative specimens, one (0.1%) was positive by the BN-LFA, resulting in a specificity of 99.9% (CI: 99.4% - 100.0%).

**Figure 2:**
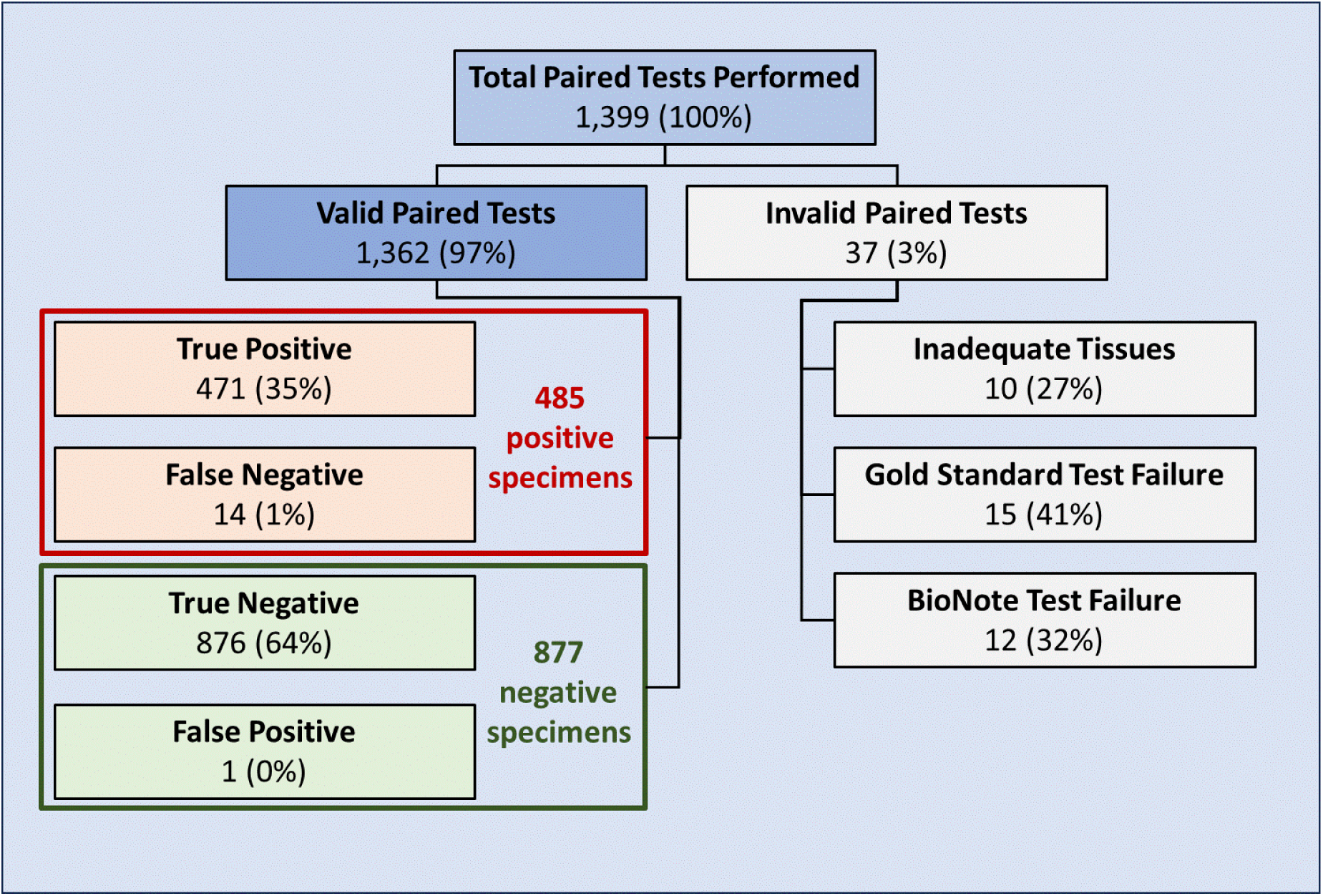
Outcomes of Paired Testing of Gold Standard Diagnostics and a Point-of-Care Test, 2024.

**Table 1:** Comparison of a BioNote Anigen Test Sensitivity and Specificity by Animal-Type, 2024.

| Animal Type | Specimens Tested | FN (n = 14) | FP (n = 1) | TN (n = 876) | TP (n = 471) | Sensitivity | Specificity |
| --- | --- | --- | --- | --- | --- | --- | --- |
| Cat | 294 | 0 | 1 | 268 | 25 | 100% | 99.6% |
| Dog | 211 | 10 | 0 | 146 | 55 | 84.6%** | 100% |
| Skunk | 205 | 0 | 0 | 80 | 125 | 100% | 100% |
| Fox | 199 | 3 | 0 | 91 | 105 | 97.2% | 100% |
| Bat | 184 | 0 | 0 | 144 | 40 | 100.0% | 100% |
| Raccoon | 100 | 1 | 0 | 45 | 54 | 98.2% | 100% |
| Horse | 21 | 0 | 0 | 16 | 5 | 100% | 100% |
| Mongoose | 21 | 0 | 0 | 0 | 21 | 100% | na |
| Bear | 16 | 0 | 0 | 16 | 0 | na | 100% |
| Bobcat | 16 | 0 | 0 | 6 | 10 | 100% | 100% |
| Coyote | 15 | 0 | 0 | 11 | 4 | 100% | 100% |
| Bovine | 13 | 0 | 0 | 2 | 11 | 100% | 100% |
| Goat | 10 | 0 | 0 | 9 | 1 | 100% | 100% |
| Other* | 57 | 0 | 0 | 42 | 15 | 100% | 100% |
| **Total** | **1,362** | **14** | **1** | **876** | **471** | **97.1%** | **99.9%** |
| US | 1,333 | 14 | 0 | 864 | 455 | 97.0% | 100% |
| International | 29 | 0 | 1 | 12 | 16 | 100% | 92.3% |

\*Moose, Wolf, Beaver, Otter, Opossum, Squirrel, Jackal, Sheep, Groundhog, Javelina, Woodchuck, Bison, Coati, Donkey, Ermine, Lynx, Mink, Wolverine \*\* When using cats (100% sensitivity) as the referent group, dogs exhibited a lower sensitivity ( $p = 0.056$ ), indicating a notable difference in performance between the two groups, though this result narrowly misses conventional thresholds for statistical significance ( $p < 0.05$ ).

Specimens from 31 animal types underwent evaluation (Table 1) [29]. Most specimens were from cats and dogs (n = 294 and 211, respectively), followed by skunks (205), foxes (199), bats (184), and raccoons (100). All other animal types had fewer than 25 specimens tested. Overall, 36% of specimens were positive by a gold standard test; higher than U.S. national averages and may reflect selective testing of known positives [29]. While 27 of the 30 animal types with positive brain tissues had 100% sensitivity compared to a gold standard test, three animal types had false-negative results with the BN-LFA: Dogs (n = 10), Foxes (n = 3), and Raccoon (n = 1). The sensitivity among these three did not significantly differ from cats, which represented the most frequently tested animal in this evaluation. However, the lower sensitivity of 84.3% for dogs narrowly missed the conventional threshold for statistical significance (p = 0.06).

Nine RVV were represented in this evaluation, including three from the inclusive bat RVV lineage (Bat, Eastern Raccoon, and South-Central Skunk RVVs) and six from inclusive dog RVV lineage (Canine, Mongoose, Arctic Fox, Arizona Gray Fox, California Skunk, and North Central Skunk) (Table 2). False-negative results from the BN-LFA were found among five RVVs; South-Central Skunk (n = 7), Arctic Fox (n = 3), North Central Skunk (n = 2), Eastern Raccoon (n = 1), and California Skunk (n = 1). The sensitivity of the BN-LFA varied from 90.9% to 100% by RVV, however, none had a significant deviation from bat-specific RVVs which was the most tested RVV group with 100% sensitivity. Dog and bat lineage sensitivity and specificity were similar (97.3% / 100% and 96.9% / 99.8%, respectively).

**Table 2:** Comparison of a BioNote Anigen Test Sensitivity and Specificity by Rabies Virus Lineage and Variant, 2024.

| Rabies Virus Variants |  | Total<br>Specimens | False-negative | False<br>Positive | True<br>Negative | True<br>Positive | Sensitivity | Specificity | Sensitivity<br>P-Value** |
| --- | --- | --- | --- | --- | --- | --- | --- | --- | --- |
| Bat Lineage<br>Variants | Bat | 199 | 0 | 0 | 156 | 43 | 100% | 100% | *referent group* |
|  | Raccoon | 393 | 1 | 0 | 227 | 165 | 99.4% | 100% | 1.00 |
|  | SC Skunk* | 267 | 7 | 0 | 179 | 81 | 92.0% | 100% | 0.09 |
|  | **Bat Lineage Total** | **859** | **8** | **0** | **562** | **289** | **97.3%** | **100%** | **na** |
| Dog Lineage Variants | Canine | 29 | 0 | 1 | 12 | 16 | 100% | 92.3% | 1.00 |
|  | Mongoose | 121 | 0 | 0 | 73 | 48 | 100% | 100% | 1.00 |
|  | Arctic Fox | 148 | 3 | 0 | 115 | 30 | 90.9% | 100% | 0.08 |
|  | Arizona Gray Fox* | 147 | 0 | 0 | 122 | 25 | 100% | 100% | 1.00 |
|  | CA Skunk | 87 | 1 | 0 | 49 | 37 | 97.4% | 100% | 0.47 |
|  | NC Skunk | 92 | 2 | 0 | 61 | 29 | 93.5% | 100% | 0.17 |
|  | **Dog Lineage Total** | **624** | **6** | **1** | **432** | **185** | **96.9%** | **99.8%** | **na** |
| *\* Arizona Gray Fox and South-Central Skunk variant territory overlaps in the state of Arizona. Negative-testing animals cannot be accurately placed into a variant category. Negative-testing foxes were placed in the Gray Fox category and negative-testing skunks were placed into the SC Skunk category. All other negative-testing animals from Arizona appear in both categories, which results in column-totals representing duplicative counting of 240 negative-testing animals. Three positive-testing animals from Arizona that were not variant typed also appear in both categories.* |  |  |  |  |  |  |  |  |  |
| *\*\* Fisher's Exact Test for sensitivity* |  |  |  |  |  |  |  |  |  |

All 14 false-negative results were found among three animal types (dogs, foxes, and raccoons) and five RVVs (Table 3). Among the 10 false-negative dogs, false-negative results occurred among samples of South-Central Skunk, California Skunk, and North Central Skunk RVVs (Table 3). All false-negative specimens from dogs occurred within U.S. laboratories. No demographic differences among false-negative dogs were observed, with specimens demonstrating a wide range of ages (1 month to 2.5 years) and breeds (Table 4). Nine false-negative dogs had no history of vaccination, and one was vaccinated shortly after rabies virus exposure. Sequencing analysis found no unique mutations among a subset of false-negative samples compared to samples that produced a positive LFA result.

**Table 3:**
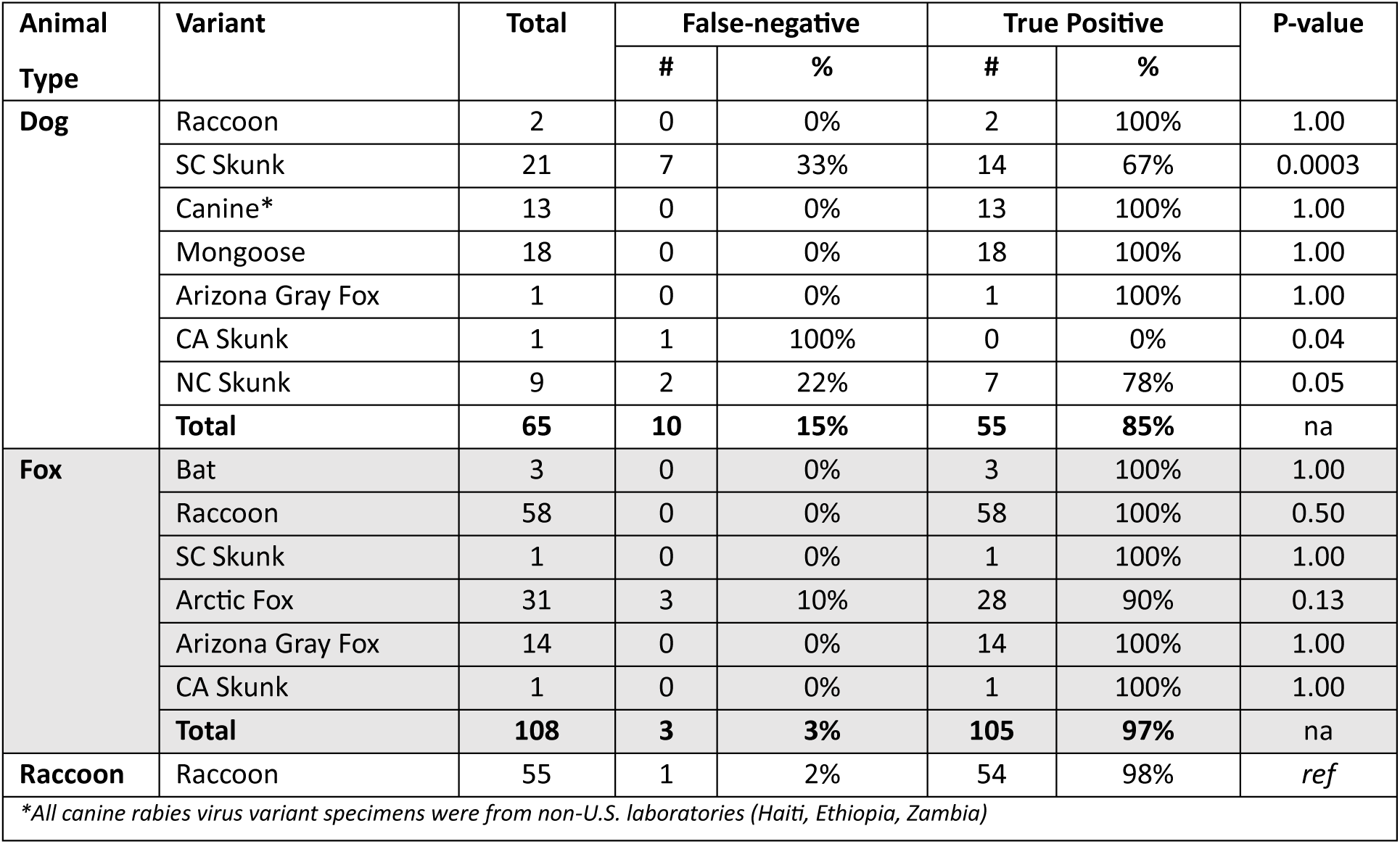
False-negative Results by Animal Type and Rabies Virus Variant, 2024.

**Table 4:** Characteristics of Rabies Positive Animals that Failed to Mount a Visible Reaction on the BioNote Anigen Test, 2024.

| Animal Type | Breed | Death Post-Symptom Onset in Days (Method) | Age | Vaccination History | Variant | Gold Standard Antigen Distribution** | Gold Standard Interpretation |
| --- | --- | --- | --- | --- | --- | --- | --- |
| Dog | Blue Heeler Mix | 7 (Natural) | 1 year | Unvaccinated | NC Skunk | 4+ | Strong |
| Dog | Yorkshire Terrier | 7 (Euthanasia) | 2.5 months | Vaccinated after exposure | SC Skunk | 4+ | Strong |
| Dog | Mixed Breed | 1 (Euthanasia) | Adult | Unvaccinated | SC Skunk | 1+ to 2+ | Weak |
| Dog | Samoyed | Unknown | 2.5 years | Unvaccinated | SC Skunk | 1+ | Weak |
| Dog | Border Collie / Australian Shephard | 1 (Euthanasia) | 1 month | Unvaccinated | SC Skunk | 2+ to 3+ | Weak |
| Dog | Husky | 1 (Euthanasia) | Juvenile | Unvaccinated | SC Skunk | 1+ | Weak |
| Dog | Bulldog Mix | Unknown | 6 months | Unvaccinated | CA Skunk | na | Unknown |
| Dog | Lab Mix | 6 (Euthanized) | 3 months | Unvaccinated | SC Skunk | 3+ | Strong |
| Dog | Boxer | Unknown (Euthanized) | 6 months | Unvaccinated | NC Skunk | 4+ | Strong |
| Dog | Unknown | 3 (Euthanized) | Puppy | Unvaccinated | SC Skunk | 3+ to 4+ | Strong |
| Red Fox |  | Unknown | na | na | Arctic Fox | CT = 31 | Weak |
| Red Fox |  | Unknown | na | na | Arctic Fox | CT = 31 | Weak |
| Red Fox |  | Unknown | na | na | Arctic Fox | 4+ | Strong |
| Raccoon |  | Unknown | na | na | Raccoon | Variable staining | Atypical |

\* No samples indicated long term storage. All had adequate sample quality.
\*\* DFA and DRIT are presented on a scale of 1 – 4, with 1 being focal distribution and 4 being wide distribution[[30, 33]

**Table 5:** BioNote Anigen Test Band Strength among Known-Positive Animals, Presented by Animal Type and Rabies Virus Variant, 2024.

| Animal / Variant |  | GS**<br>Positive<br>Specimens | Strong |  | Normal |  | Weak |  | No Band |  | Weak or No Band |  |
| --- | --- | --- | --- | --- | --- | --- | --- | --- | --- | --- | --- | --- |
|  |  |  | n | % | n | % | n | % | n | % | n | % |
| Animal Type | Skunk | 125 | 28 | 22% | 94 | 75% | 3 | 2% | 0 | 0% | 3 | 2% |
|  | Fox | 108 | 50 | 46% | 52 | 48% | 3 | 3% | 3 | 3% | 6 | 6% |
|  | Raccoon | 55 | 35 | 64% | 10 | 18% | 9 | 16% | 1 | 2% | 10 | 18% |
|  | Dog | 65 | 3 | 5% | 42 | 65% | 10 | 15% | 10 | 15% | 20 | 31%** |
|  | Bat | 40 | 7 | 18% | 28 | 70% | 5 | 13% | 0 | 0% | 5 | 13% |
|  | Cat | 25 | 12 | 48% | 11 | 44% | 2 | 8% | 0 | 0% | 2 | 8% |
|  | Mongoose | 21 | 3 | 14% | 17 | 81% | 1 | 5% | 0 | 0% | 1 | 5% |
|  | Bovine | 11 | 4 | 36% | 1 | 9% | 6 | 55% | 0 | 0% | 6 | 55%*** |
|  | Bobcat | 10 | 3 | 30% | 6 | 60% | 1 | 10% | 0 | 0% | 1 | 10% |
|  | Other* | 25 | 7 | 28% | 14 | 56% | 4 | 16% | 0 | 0% | 4 | 16% |
| Rabies Virus Variant | Raccoon | 166 | 108 | 65% | 38 | 23% | 19 | 11% | 1 | 1% | 20 | 12% |
|  | SC Skunk | 85 | 22 | 26% | 52 | 61% | 4 | 5% | 7 | 8% | 11 | 13% |
|  | Mongoose | 48 | 5 | 10% | 38 | 79% | 5 | 10% | 0 | 0% | 5 | 10% |
|  | Bat | 43 | 7 | 16% | 29 | 67% | 7 | 16% | 0 | 0% | 7 | 16% |
|  | CA Skunk | 41 | 0 | 0% | 38 | 93% | 2 | 5% | 1 | 2% | 3 | 7% |
|  | Arctic Fox | 33 | 4 | 12% | 26 | 79% | 0 | 0% | 3 | 9% | 3 | 9% |
|  | NC Skunk | 31 | 2 | 6% | 20 | 65% | 7 | 23% | 2 | 6% | 9 | 29%** |
|  | Arizona Gray | 22 | 3 | 14% | 19 | 86% | 0 | 0% | 0 | 0% | 0 | 0% |
|  | Canine | 16 | 1 | 6% | 15 | 94% | 0 | 0% | 0 | 0% | 0 | 0% |
| *\*Bison, Coyote, Donkey, Goat, Groundhog, Horse, Jackal, Moose, Opossum, Otter, Woodchuck*<br>*\*\*GS = Gold Standard (Tests include Direct fluorescent Antibody (DFA), Direct Rapid Immunohistochemistry (DRIT) and LN34 Real-time RT-PCR.)*<br>*\*\*\* Value is outside of the 95% confidence interval for either Animal Type or Rabies Virus Variant.* |  |  |  |  |  |  |  |  |  |  |  |  |

Among 485 positive specimens in this evaluation, 16% had weak or false-negative test bands (Table5). Rabid dogs and bovines had significantly higher proportions of tests demonstrating weak or false-negative test bands (31% and 55% of positive specimens, respectively). When stratified by RVV, 11% of specimens had weak or false-negative test bands. Only the North Central Skunk variant had a significantly higher rate, with 29% displaying a weak or false-negative test band. When the test band was described as strong or normal, only 1.2% of specimens had a weak gold standard reaction (three of 413 specimens). Among 44 tests that reported a visible but weak test band, three were also notably weak by the gold standard (7%). Among the 14 false-negative results where no positive band was visible, eight had a weak gold standard test reaction (57%).

The LN34 assay produced Ct values below the diagnostic threshold for dilutions up to -2 (1:100) in the Caribbean Mongoose RVV series, up to -4 (1:10,000) in the Eastern Raccoon RVV series, and -5 (1:100,000) in the South-Central Skunk RVV series. The LN34 Assay produced a Ct value of 36 in Eastern Raccoon RVV dilution -5, which is above the diagnostic threshold of 35. The LN34 assay did not produce Ct values for dilutions -3, -4, and -5 in the Caribbean Mongoose RVV series. The LFA produced a visible positive band for the undilute sample for the Caribbean Mongoose RVV, dilutions up to the -2 (1:100) for both the Eastern Raccoon RVV, and South-Central Skunk RVV series. No differences were noticed between the BN-LFA with Eastern Raccoon RVV and South-Central Skunk RVV. These results indicate that LN34 is approximately 100 times more sensitive than the BN-LFA. Samples with a Ct value 23 to 27 are at higher risk of a false-negative reaction, and 27 or higher would likely result in false-negative interpretation (Figure 3).

**Figure 3:**
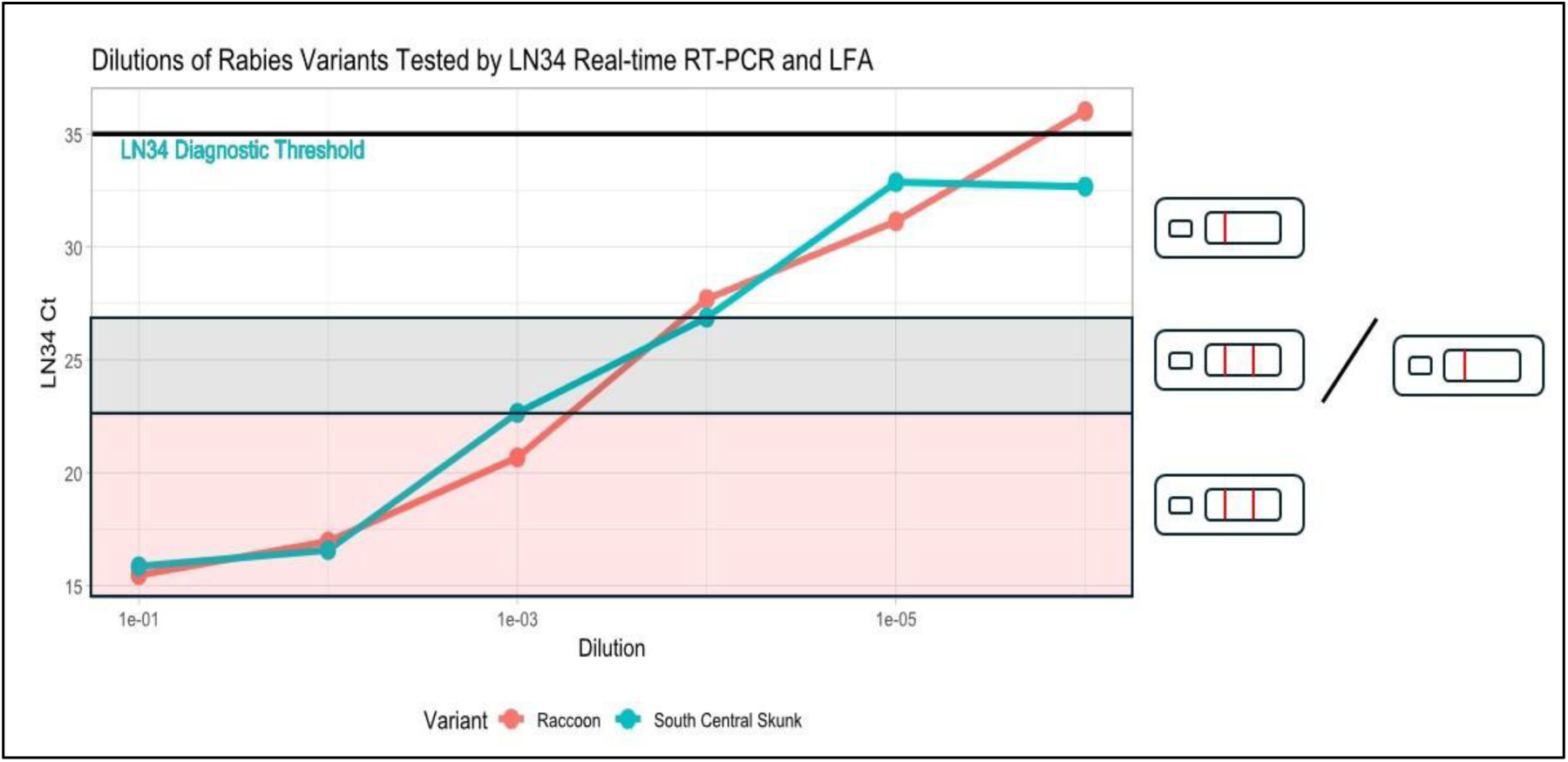
Comparison LN34 and LFA of RVV Test Detection Limits. *\*Red shading indicates likely high concordance between the BioNote Anigen Test and the LN34 PCR, when the LN34 CT value is below 23. Gray shading indicates uncertain agreement, when the LN34 CT value is between 23 and 27. BioNote Anigen Test is likely to result in a false-negative result when the CT value is above 27 (white shading)*.

## Discussion

Point-of-care tests for rabies diagnosis have long been sought-after for improving rabies surveillance in low- and middle-income countries [13, 38, 40], yet all prior large-scale evaluations have found significant faults in commercially available test kits [20, 41]. Rabies test results are often used to inform human life-saving vaccination decisions and direct expensive response actions [42, 43]. In 2023, WOAH cautioned against the use of unvalidated LFA tests outside of research or evaluation programs [14]. Despite this statement, numerous field deployments of these tests have been published, demonstrating the high demand for a diagnostic method that overcomes barriers of current gold-standard methods [15-26].

This is the largest to-date evaluation of a point-of-care test for rabies and the first to assess the BN-LFA against diverse rabies virus variants. CDC maintains laboratory and epidemiologic expertise for the development, validation, and implementation of diagnostic assays. All laboratories included in this study received CDC training and represent the capacities of national reference laboratories found in many countries. The U.S. is endemic for >13 known bat-maintained RVVs and seven terrestrial-maintained RVVs, including the Mongoose RVV found only in Puerto Rico, and which is antigenically indistinguishable from its predecessor cosmopolitan dog-maintained variant. Additionally, CDC-trained laboratories in Haiti, Ethiopia, and Zambia provided specimens representing dog-maintained RVVs.

Overall, the BN-LFA performed well with sensitivity and specificity results that are comparable to globally recognized gold standard tests such as the DFA and DRIT [6, 8]. However, this evaluation demonstrates that false-negative reactions are possible and likely to occur at very low rates. Extensive efforts were undertaken to understand the cause of these BN-LFA failures. False-negative samples were clustered among U.S.-origin dogs but had no clear predilection for a specific RVV or species-RVV combination. Random error and statistical significance were considered, however, this study boasts a large sample size and several statistical measures of significance were identified, particularly when dogs were infected with any of the three genetically and antigenically diverse U.S. skunk RVV (California Skunk, North Central Skunk and South-Central Skunk RVV) [44].

Host-virus interactions were considered but deemed unlikely to have contributed to the false-negatives in this evaluation. False-negative samples were found across five different RVVs and two different viral lineages, representing diverse viral genetics that have no common point-mutation that would infer weakened antibody binding [44]. Further, sequencing analysis of a subset of the false-negative samples found no mutations likely to impact antibody binding; only very minimal genetic differences of several amino acids were noted between these samples and those that produced a strong reaction. While theoretically plausible, host pressure on the rabies virus is unlikely to result in significant mutations due to the relatively conserved viral genome and characteristically low mutation rate [44]. Additionally, sequencing results and adequate reactions produced by gold standard test methods do not seem to support any meaningful *in vivo* viral genetic changes that should lead to test failure.

Host-factors among rabid animals that failed to produce a visible band on the BN-LFA were also investigated, particularly among the 10 false-negative dogs. Very few similarities were noted among these dogs, with failures represented among a broad range of ages, breeds, geographic locations, testing laboratories, and RVVs. Theoretically, a history of vaccination could result in host-produced neutralizing antibody, which if serum or blood were introduced to the specimen, could bind virus prior to application on the device thereby producing false-negative reactions [45]. However, nine of these dogs had no history of vaccination. Rabies antibody is rarely produced prior to rabies virus symptom onset, and many mammals die before meaningful antibody is produced in the serum or CNS, therefore this was almost certainly not a factor leading to the false-negative results [46].

Interestingly, most false-negative dogs were euthanized relatively early in the course of disease, a common practice in upper income countries when rabies is suspected or successful treatment outcomes are deemed unlikely [47, 48]. Whereas wildlife species and dogs in resource-limited settings are often found dead or euthanized relatively late in the clinical phase of disease. This may suggest that viral load is associated with false-negative test results [49]. Supporting this theory, there was a direct association between the strength of the BN-LFA’s positive band and the gold standard test results reaction, with over 50% of false-negative tests having weak gold standard reactions. Further supporting the theory that low viral loads are associated with LFA false-negative results, the LN34 real-time RT-PCR demonstrated at least 1:100 times more sensitive results than the BN-LFA. These results indicate that 100 times more viral load is required to obtain a positive result by the BN-LFA than the gold standard LN34 assay.

While viral load in the specimen is almost certainly a primary factor leading to false-negative test results for the BN-LFA, there were several specimens which had notably strong gold standard reactions but still were reported as non-visible LFA reactions. While CDC-trained, high-quality laboratories were selected for this evaluation, point-of-care tests rely on a colorimetric reaction that must be readily visible to the device operator. As noted by the device manufacturer, any colorimetric change, regardless of how faint, should be considered a positive reaction. There is also a time-component to this test, with a positive reaction occurring up to ten minutes after the sample is deposited onto the device. In this evaluation, 12% of BN-LFA reactions were notably weak or absent in true positive samples. While low viral load can directly result in a false-negative result, additional factors such as operator attentiveness to a faint band reaction or reading the device too early may have contributed to the rare false-negative results identified in this evaluation.

Gold standard antigen detection tests such as DFA use two anti-rabies conjugates in each test, each conjugate containing two or three pan-reactive monoclonal antibodies to ensure broad detection of RVVs [8, 30, 42]. The BN-LFA could increase the monoclonal antibody concentration within the test cassette and use more than one pan-reactive monoclonal antibody to increase the potential for low antigen recognition. While the BN-LFA worked well for all RVVs in this evaluation, lack of clarity on the antibodies used in this test raise concerns for broad assumptions that the test will work across all RVVs [50, 51].

Past evaluations of commercially available rabies test kits found concerningly high rates of lot-to-lot variation [20, 41]. Five BN-LFA lots were included in this study, and periodic checks of sensitivity and specificity demonstrated that all had equal performance. This may suggest an improvement in production quality of the devices compared to past evaluations. Commercial test kits are sensitive to changes in manufacturing practices. Reagents for most diagnostic tests undergo external validation prior to distribution of the lots [52]. If the BN-LFA is to be WOAH/WHO recognized test for rabies diagnosis, a similar lot-to-lot validation process with rabies reference laboratories should be considered to ensure consistent device quality.

This evaluation found a high sensitivity and specificity for the BN-LFA, with values similar to those reported of other gold standard rabies tests [6, 8]. While false-negatives were identified, this should be expected for any rapid antigen detection methods, particularly when compared to extremely sensitive molecular diagnostic methods. The clustering of false-negatives among dogs, one of the intended species for use of this test, is most likely an artifact of early euthanasia commonly practiced in North America and rare occurrences of operator error. This evaluation focused on the predominant terrestrial and bat RVVs endemic in the USA, these highly divergent viruses were not well represented in previous commercial LFA studies and when previously tested, demonstrated unacceptable sensitivities (40-60%). As with any new diagnostic method, evaluation studies should continue to be performed, as well as evaluation of this test’s performance across the 16 other lyssaviruses and any RVV in the programmatic areas they are deployed [42, 53]. The results obtained within this study are based on the modifications to the original BioNote instructions. Collection of a complete cross-section of brain stem thorough homogenization to a paste, careful coating of the swab and blending into the diluent buffer are all essential to the significant increase in test performance observed in this study. Despite the rare failures of the BN-LFA, there is clearly a high and well-deserving demand for rabies rapid antigen tests. If treatment and programmatic response decisions are paired with veterinary assessments of clinically suspicious animals and confirmatory testing with a gold standard assay (DFA, DRIT and LN34 assay), negative veterinary or human health outcomes stemming from widescale use of the BN-LFA are likely negligible.

## Data Availability

All data produced in the present study are available upon reasonable request to the authors

## Supplementary Figures and Tables

### Appendix A

**BIONOTE Rabies Anigen Test Kit® Study Comparison of Lateral Flow Assay (LFA) (CDC Quick Reference Protocol)and Direct Fluorescent Antibody Test [30]** provided to all participating laboratories

### Introduction

#### General safety

All rabies diagnostic procedures should be performed following biosafety precautions for handling Lyssaviruses and following the laboratory’s site-specific risk assessment including review of the relevant best practices in the current Biosafety in Microbiological and Biomedical Laboratories- 6^th^ Edition (BMBL). Prior to start of testing, risk assessments should account for the vaccination status of laboratory personnel providing testing (pre-exposure rabies immunization and monitoring of titer), as well as the protocols (brain collection, test protocols, waste disposal and decontamination) and the PPE required at each step. The minimum personal protective equipment (PPE) required for Lateral Flow Assay (LFA) testing after brain removal includes: lab coat, safety glasses, double gloves and N95 mask; face shields are optional. These biosafety precautions adequately address all reasonable rabies virus exposure concerns. If a BSC is used, then N95 mask, safety glasses and face shield are not required.) If there are variations in these recommendations, additional steps can be taken to mitigate the risk of rabies virus exposure; a site-specific risk assessment should be performed to determine additional mitigations that may be necessary.

All reusable materials and equipment used in testing should be decontaminated using a recommended disinfectant (1:256 QAC, 10% bleach, 10% iodophor) after use, and disposable materials should be incinerated or autoclaved before discard. Personnel should be aware of the potential hazards and how to avoid or reduce these hazards prior to initiating rabies testing (WHO Laboratory Techniques in Rabies Chapter Biosafety 2018, BMBL 2020). If the Lateral Flow Assay is to be performed in a laboratory and a Class II BSC is available, then it should be used for this testing. However, the BioNote Rabies Anigen Test Kit was designed to be a rapid field test and can be performed easily and safely if the aforementioned biosafety recommendations are taken. If this protocol is modified in a manner that may result in aerosolization of rabies virus, additional biosafety actions should be considered.

#### Principle of the test

Lateral flow assays, which are more formally known as rabies immunochromatographic diagnostic tests (RIDC), are most frequently used to detect rabies virus antigen. A brain suspension is added to the opening in the device cassette and is adsorbed into the sample pad (Supplemental Figure 1). The sample then flows through the conjugate pad containing anti-rabies antibodies labeled with gold nanoparticles.

**Supplemental Figure 1.**
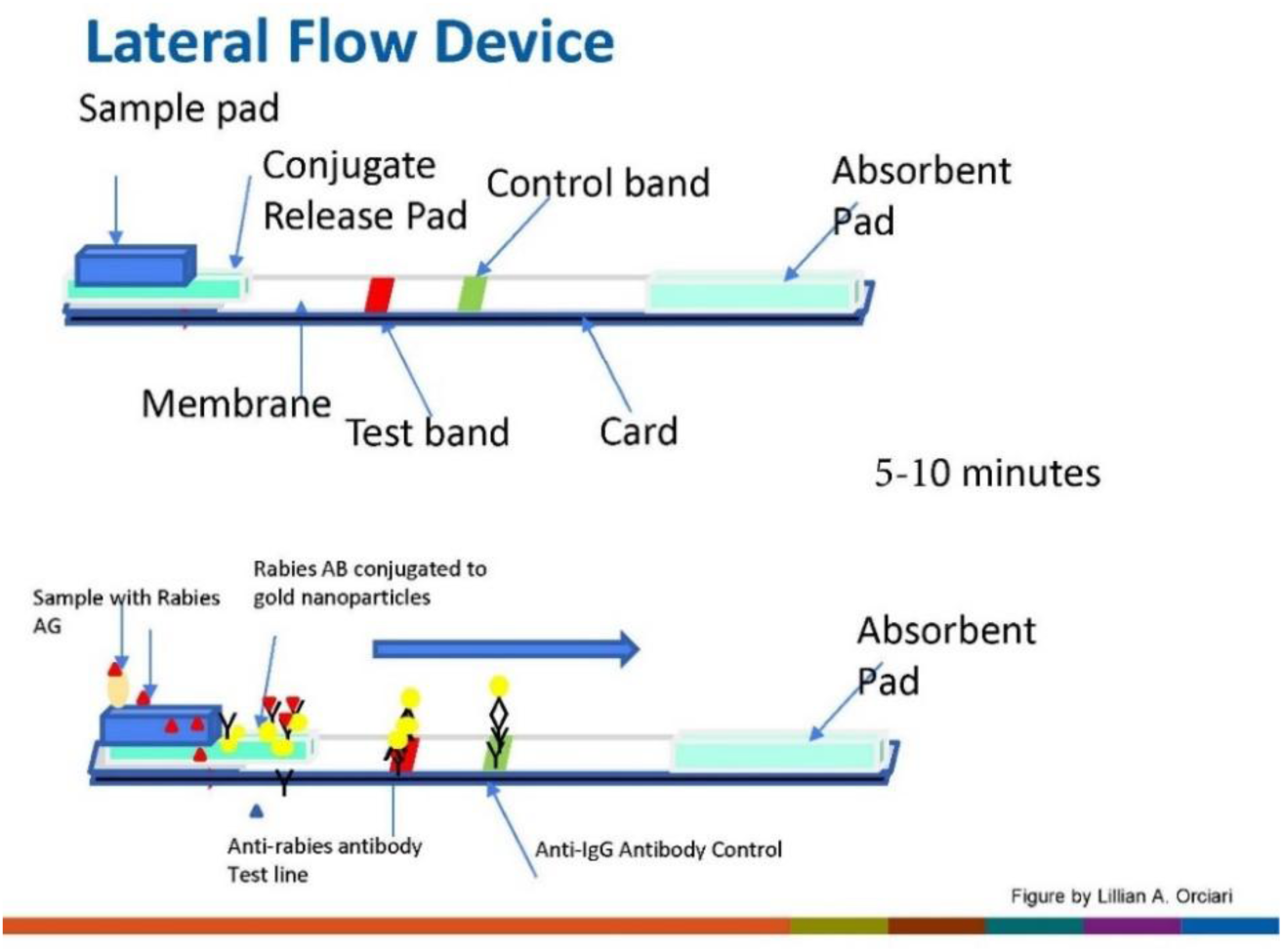

If rabies virus antigen is present, nanoparticle-labeled antigen-antibody complex diffuses down the membrane strip and reacts with the test band containing a second anti-rabies antibody and a colored band appears (Supplemental Figure 2). When the suspension flows to the control band, the anti-IgG will react with the anti-rabies antibodies and the control band will appear. For the test to be valid, the reagents would need to flow appropriately along the membrane strip, and the conjugate labeled with nanoparticles (either bound with antigen or unbound in case of a negative sample) must react with the anti-IgG control band.

**Supplemental Figure 2.**
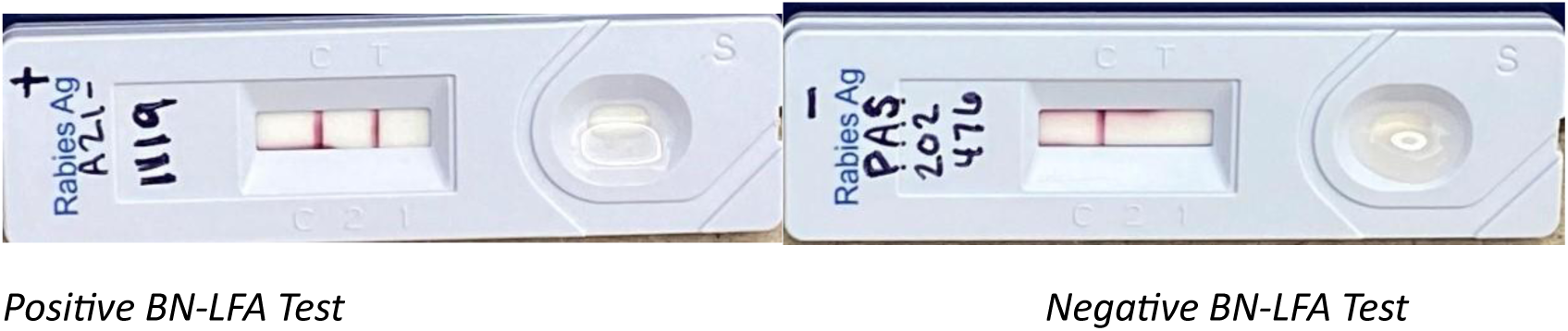

While the concept of these assays is good, in reality LFAs have not performed as well as expected (lower sensitivity and specificity) when used according to the manufacturers’ recommendations. CDC is currently working on a method to improve the sensitivity of the LFAs for rabies antigen detection and will be working with the collaborators in the USA as well as international partners to test more thoroughly with the modifications proposed here.

### Brain Collection

The brain sample (brain stem, full cross-section) is collected for the LFA using the same standard operating procedure and PPE as for collection of brain stem for DFA or DRIT. This sample can be taken from dorsal side or ventral side via the foramen magnum using a scalpel or spatula as well as from a fully excised brain sample. For this study, a full cross-section of brain stem must be tested, and sufficient tissue collected for LFA, DFA or DRIT, and confirmatory testing at a reference laboratory in the case of discordant findings. To ensure the identical cross-section of brain stem is tested by both LFA and DFA or DRIT, harvest a single cross-sectional piece of brain stem for the tests required and place it in a container so that the orientation can be easily identified. With care, immediately make a cross-sectional slice through the brain stem piece for LFA and place it in a separate container. Repeat this process making adjacent cross-sectional slices for DFA or DRIT and confirmatory testing.

#### DO NOT USE THE STRAW COLLECTION METHOD

Brain tissues cannot be easily identified when extracted using the straw, especially tubular pieces extracted of neocortex (white tissues from the cerebral cortex) cannot be easily distinguished from brain stem. Depending on the size of the animal, the straw method may not provide full cross-section of the brain stem, impacting interpretability of the test.

### LFA method modified by CDC

The BioNote “Rabies Anigen Test Kit” was designed to be a rapid field test and can be performed easily and safely in the field providing additional containment steps are performed to avoid or reduce potential risks. If the test can be performed in a laboratory with a BSC then the additional containment options are not necessary. If the aforementioned biosafety measures are followed, no additional biocontainment is necessary for using the LFA in the field (Option 1). If there are deviations in the biosafety procedures that may result in aerosolization of brain homogenate, then additional biocontainment options should be considered (Option 2).

Option 1: Part of the procedure is performed outside of the bag.

Option 2: The entire LFA protocol may be performed inside of a 1 gallon size (12”x12”x12”) plastic ziplock bag (Supplemental Figure 3).

**Supplemental Figure 3.**
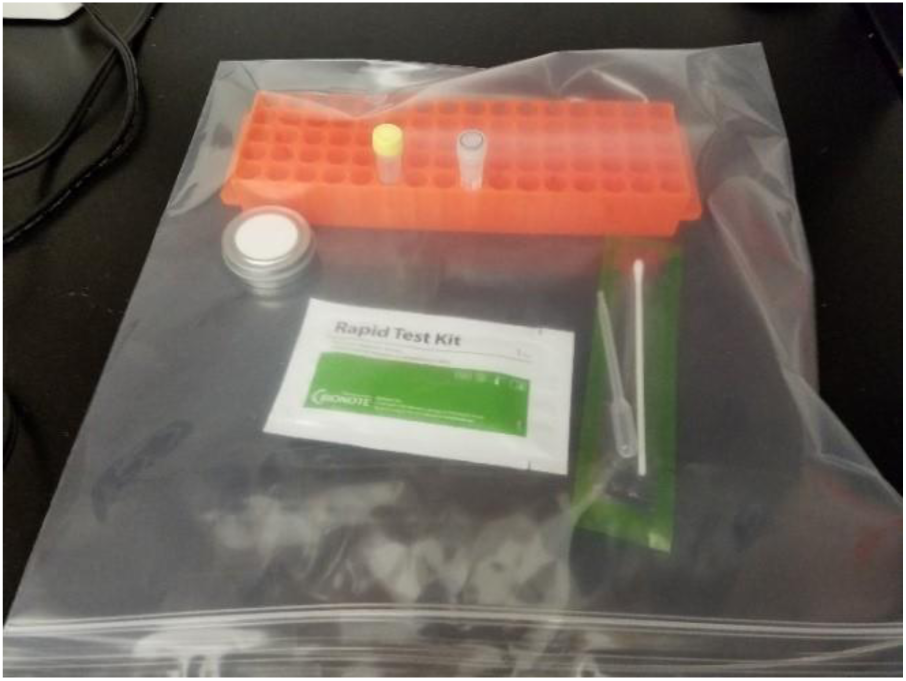

#### Materials needed (Not provided in the BioNote Test Kit)

Gallon size ziplock bags (optional, based on biosafety assessment)

2 ml Sarstedt tubes (screw cap tubes equiped

Autoclavable plastic test tube rack for 2 ml Sarstedt tubes

Sharps container for scalpels and other sharps

Scalpels

Discard container with disinfectant (e.g. 1:256 QAC, 10% bleach, 10% iodophor)

PPE (lab coat, N95 mask, gloves, safety glasses and/or faceshield)

Permanent fine tip marker

Lab mat

Autoclave bag for disposable material

**Materials needed** (Provided in the BioNote Test)

Anigen Rapid Rabies Ag Test Devices

Assay diluent tubes

Disposable swabs

Disposable droppers

### CDC SOP

Allow reagents to come to room temperature prior to use.

If a BSC is not available or biosafety recommendations cannot be fully adhered to, place a lab mat on a level bench surface and a gallon size ziplock bag containing, the test tube rack with an empty Sarstedt tube and assay diluent tube, 1 swab and Anigen Rapid Rabies Ag Test Device. Within the BSC or inside large Ziplock bag arrange the needed materials (Supplemental Figure 3 from previous page).

1. Take a piece of brain stem (full cross-section) usually (0.1 g to 0.2 g), the size of the (BioNote) cotton tip (Supplemental Figure 4). If the brain stem is from a large animal the full cross section should still be collected.
2. Mince tissue as finely as possible (to paste Supplemental Figure 5a) with a scalpel (best to have tin securely on the bench while cutting tissue to avoid potential sharps injury with scalpel (held in photo to get best angle for demonstration and pictures) and transfer the tissue to 2 ml Sarstedt tube or 15 ml centrifuge tube for large animals used for coating the swab (Supplemental Figure 5b). It would be safer and preferrable to transfer the minced tissue to the tube with a swab rather than the scalpel as shown here.
3. With the swab provided in the kit (Supplemental Figure 6), rub the brain tissue against the inside of the tube with the swab until the brain consistency is a smooth paste and the swab is coated with brain (Figure 6). This process may up to 1 minute or more depending on how well the brain was minced in the previous step. The tube should face away from any personnel in the vicinity, to avoid unintentional splashes.
4. Transfer the swab coated with the brain from the Sarstedt tube to the assay diluent tube. (There should be no chunks of tissue, resulting from brain that was not fully minced to a paste).
5. Prepare an even suspension in the assay diluent tube by rubbing the swab coated with brain paste gently against the inside of the tube for <u>at least</u> 10 seconds until brain is completely blended into the diluent, it may take longer (*Supplemental Figure 7*). *Supplemental Figure 7 (note, use of Ziploc bag is optional, based on adherence to recommended biosafety measures for field use when a BSC is not available)*.
6. If a bag is used, carefully open just prior to applying the sample. Label the LFA with a permanent marker (date, sample #, initials of operator).
7. Add 4 drops of the sample to the well on the LFA with the disposable dropper provided in the BioNote kit. Set timer for 10 minutes (*Supplemental Figure 8*). *Supplemental Figure 8 (note, use of Ziploc bag is optional, based on adherence to recommended biosafety measures for field use when a BSC is unavailable)*.
8. Movement of the sample liquid into the secondary chamber of the LFA should be noticed in about one minute. If movement of sample is not noticed add another drop (<u>only one</u>) of the sample (Figure 9). *Supplemental Figure 9 (note, use of Ziploc bag is optional, based on adherence to recommended biosafety measures for field use)*.
9. Record results (on the device and worksheet) once the band(s) are apparent in 5-10 minutes. After 10 minutes the reactions are complete and final. Reading results after 10 minutes could potentially lead to false positive results (Figure 10), however it has not been noticed in these studies. *LFA Demo sample is negative after 10 minutes (Control band and no test band)*
10. Take a photograph at 10 minutes incubation for a permanent record.

**Supplemental Figure 4.**
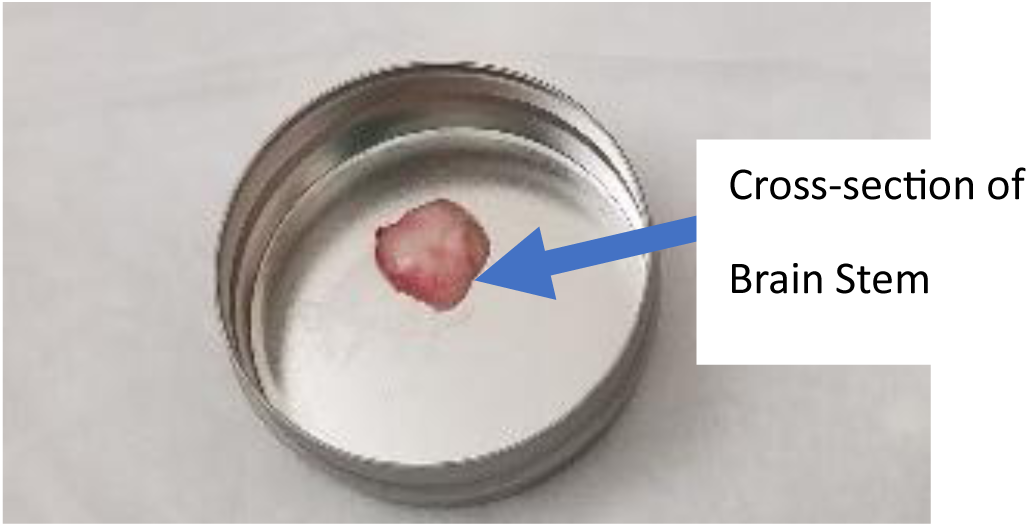

**Supplemental Figure 5 (a and b).**
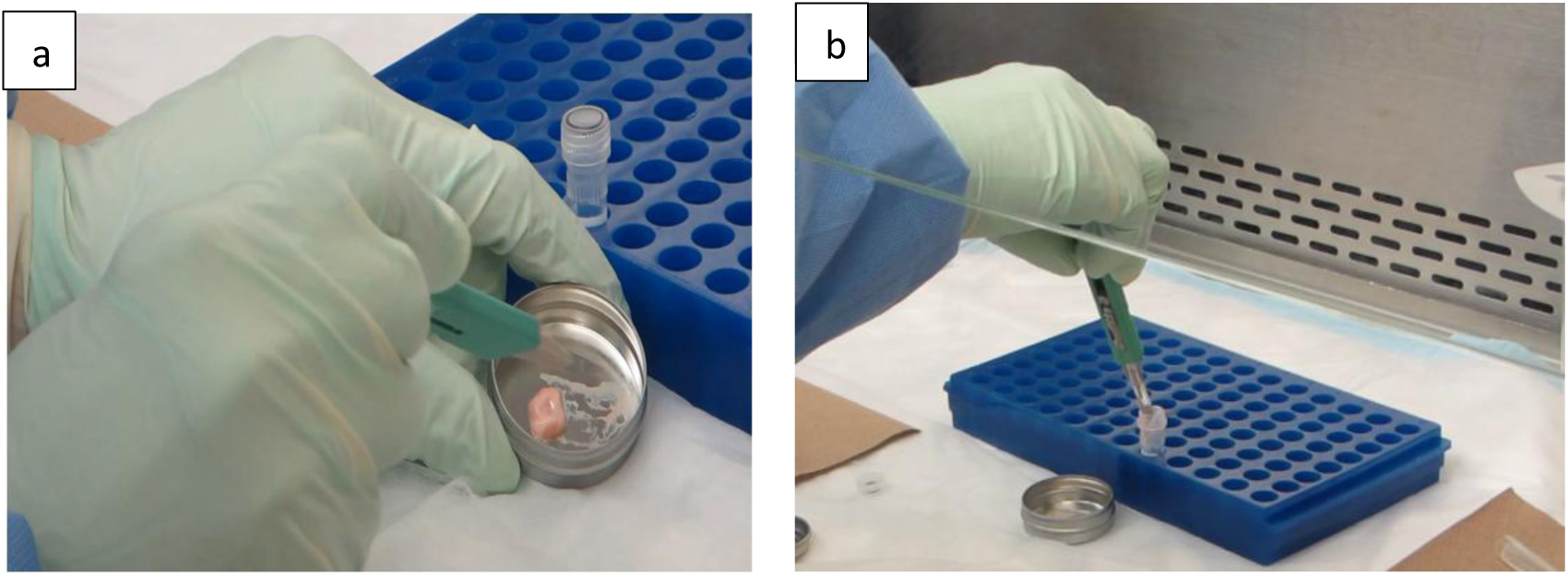

**Supplemental Figure 6.**
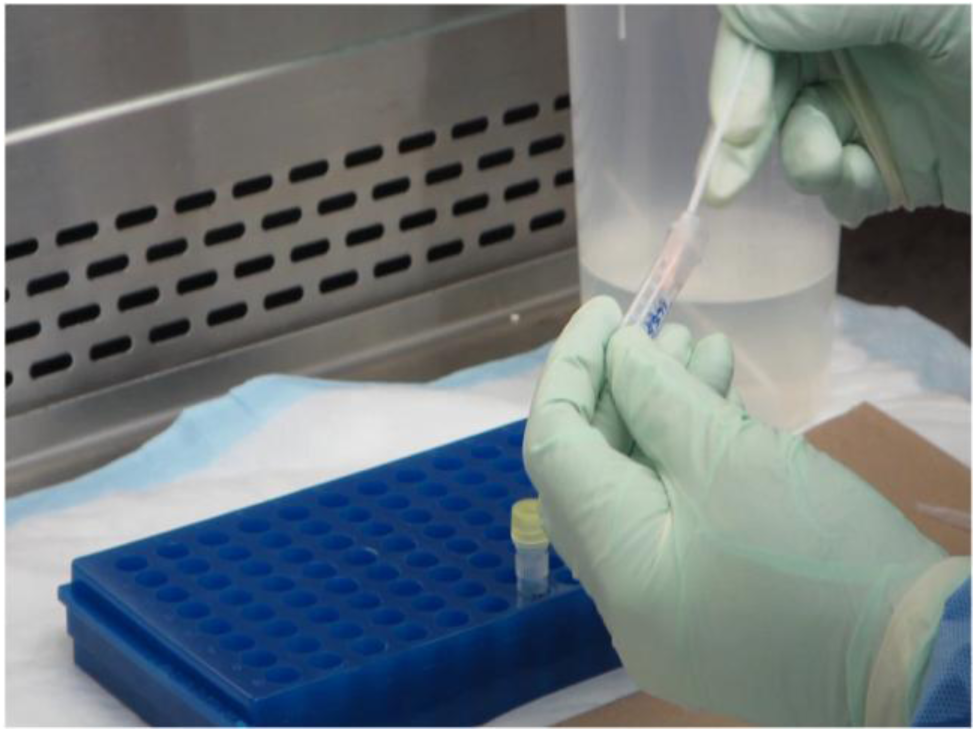

**Supplemental Figure 7.**
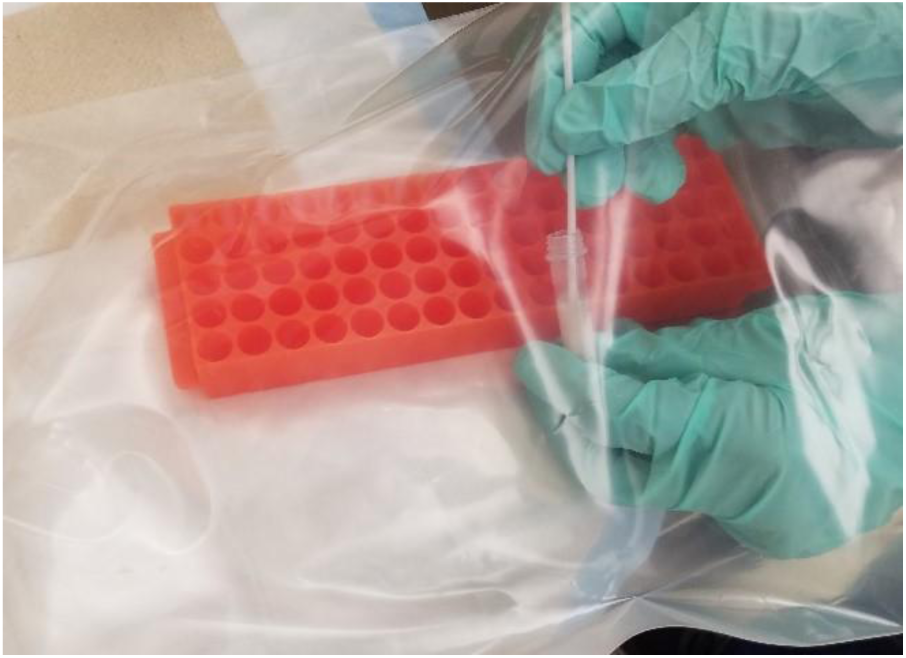

**Supplemental Figure 8.**
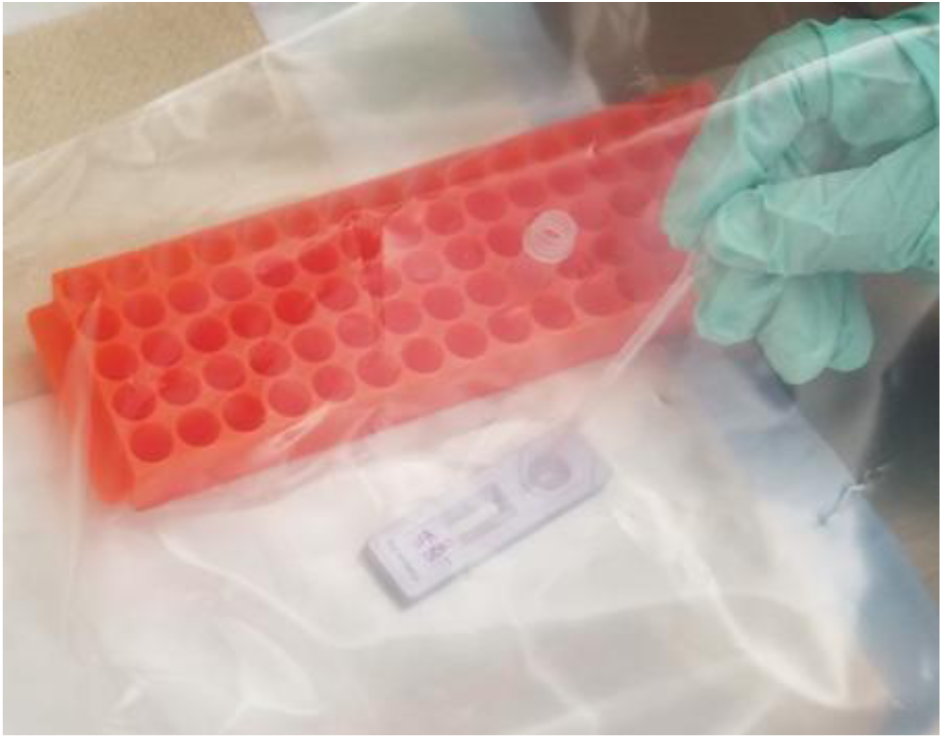

**Supplemental Figure 9.**
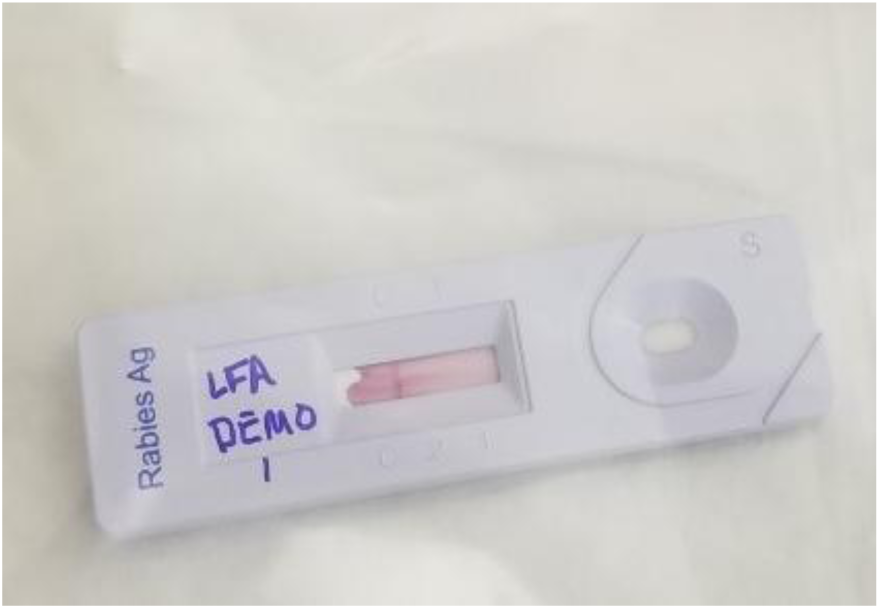

**Supplemental Figure 10.**
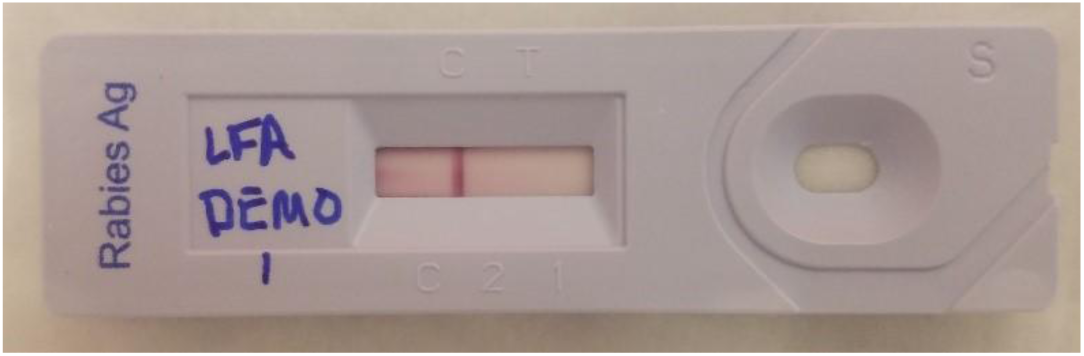

#### Interpretation of results

11. Negative results: C (Control) band only, Positive results C and T (test) bands present, and invalid results no C band present (*Supplemental Figure 11*).
12. Invalid results should be repeated immediately using another LFA kit. If repeat test is invalid, test will be reported as invalid.
13. Discordant results with the DFA or DRIT can also be repeated using this same procedure. Do not replace the original results in the tracking sheet, rather start a new record that clearly denotes that this is a repeat test based on discordant results.

**Supplemental Figure 11.**
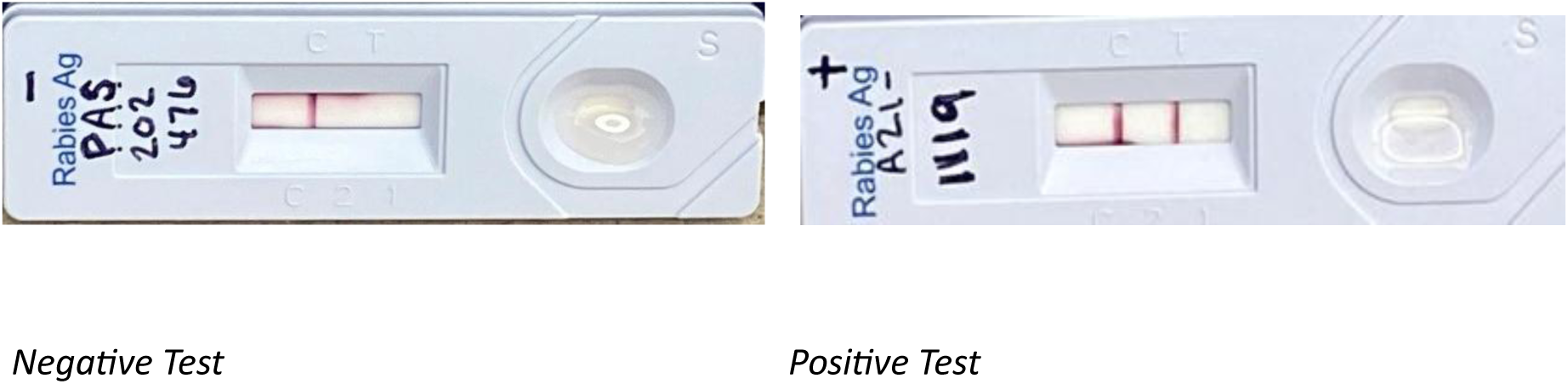
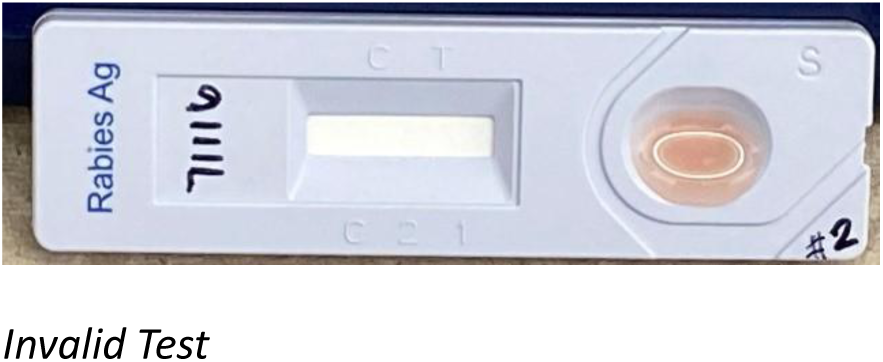

#### Decontamination and clean up

14. Tubes swabs and LFA can be placed in a container of appropriate disinfectant and autoclaved, lab mat and other disposable material may be autoclaved or incinerated prior discard.
15. Work surfaces and non-disposable equipment used for testing should be decontaminated using an approved disinfectant after testing is completed.

## Acknowledgements

Lauren Hovis^1^,

Hilary Tamnanchit^4^

Aimee Rendon^4^

Robert DeLeon^4^

Yocelyn Cruz^4^

Santana Mendoza^5^

Alexis Emerson^5^

Uyen-Kim Le^5^

Wittnee Estes^5^

Mickaela Newsum^5^

Ksenia Poujlivaia^5^

Uyen-Kim Le^5^

Wittnee Estes^5^

Mickaela Newsum^5^

Ksenia Poujlivaia^5^

Camilla Lieske^6^

Lindsey Dreese^6^

Amy Liefer^8^,

Matthew Gunkel^8^

A. Mason^9^

M. Ray^9^,

S. Buch^9^

Nachea Qualls

Rabies Identification Team, Texas Dept of State Health Services, Public Health Laboratory Division 1100 W 49^th^ St. MC 1947, Austin, TX 78756 ^12^

## Standard Disclaimer

The findings and conclusions in this report are those of the authors and do not necessarily represent the official position of the Centers for Disease Control and Prevention.

